# Strategies to support substance use disorder care transitions from acute-care to community-based settings: A Scoping review and typology

**DOI:** 10.1101/2023.04.24.23289042

**Authors:** Noa Krawczyk, Bianca D. Rivera, Ji E. Chang, Margaux Grivel, Yu-Heng Chen, Suhas Nagappala, Honora Englander, Jennifer McNeely

**Affiliations:** Department of Population Health, NYU Grossman School of Medicine, New York NY 10065; Department of Public Health Policy and Management, NYU School of Global Public Health, New York NY 10003; Department of Social and Behavioral Sciences, NYU School of Global Public Health, New York NY 10003; Department of Criminal Justice, Temple University, Philadelphia, PA 19102; University of California Berkeley, Berkeley, CA 94720; Department of Medicine, Oregon Health & Science University, Portland, OR 97239

**Keywords:** Care transitions, care navigation, warm handoff, substance use disorder, treatment, hospital, emergency department, acute-care, interventions, opioid use disorder

## Abstract

**Background:** Acute-care interventions that identify patients with substance use disorders (SUDs), initiate treatment, and link patients to community-based services, have proliferated in recent years. Yet, much is unknown about the specific strategies being used to support continuity of care from emergency department (ED) or inpatient hospital settings to community-based SUD treatment. In this scoping review, we synthesize the existing literature on patient transition interventions, and form an initial typology of reported strategies.

**Methods:** We searched Pubmed, Embase, CINAHL and PsychINFO for peer-reviewed articles published between 2000-2021 that studied interventions linking SUD patients from ED or inpatient hospital settings to community-based SUD services. Eligible articles measured at least one post-discharge treatment outcome and included a description of the strategy used to promote linkage to community care. Detailed information was extracted on the components of the transition strategies and a thematic coding process was used to categorize strategies into a typology based on shared characteristics. Facilitators and barriers to transitions of care were synthesized using the Consolidated Framework for Implementation Research.

**Results:** Forty-five articles met inclusion criteria. 62% included ED interventions and 44% inpatient interventions. The majority focused on patients with opioid (71%) followed by alcohol (31%) use disorder. The transition strategies reported across studies were heterogeneous and often not well described. An initial typology of ten transition strategies, including five pre- and five post-discharge transition strategies is proposed. The most common strategy was scheduling an appointment with a community-based treatment provider prior to discharge. A range of facilitators and barriers were described, which can inform efforts to improve hospital-to-community transitions of care.

**Conclusions:** Strategies to support transitions from acute-care to community-based SUD services, although critical for ensuring continuity of care, vary greatly across interventions and are inconsistently measured and described. More research is needed to classify SUD care transition strategies, understand their components, and explore which lead to the best patient outcomes.

## Introduction

In the midst of an ongoing drug overdose crisis, engaging individuals with substance use disorders (SUDs) in evidence-based treatment and services is an urgent priority. And yet, many at-risk patient populations with SUD remain largely disconnected from care[1–3]. Opioids remain the leading cause of overdose death, but an estimated 87% of individuals with opioid use disorder (OUD) do not access evidence-based treatment with methadone or buprenorphine[4].

One approach to address the gap in OUD treatment, and SUD treatment more broadly, has been to leverage emergency department (ED) and inpatient hospital encounters as potential “touchpoints” to offer patients with SUD an opportunity to initiate treatment (including medications for opioid use disorder (MOUD), when appropriate) and link them with ongoing services in the community[5]. Indeed, a growing body of literature is emerging regarding both ED [6] and inpatient hospital models [7] to deliver care for SUD patients. These practices generally involve screening and assessing patients for unhealthy substance use (hazardous use or SUD), offering a brief intervention and/or initiation of pharmacotherapy, and referring patients to ongoing treatment upon discharge [8–10] The evidence on the effectiveness of these interventions in improving linkage to treatment following hospital discharge has been mixed [11], with interventions offering initiation of MOUD for OUD showing more promise in recent years [12,13].

While SUD interventions have proliferated across acute-care settings, much is still unknown about the particular practices used to support the transition of patients from the hospital to community-based care. A recent systematic review of interventions to support SUD patients upon hospital discharge found great heterogeneity in the settings, approach, and nature of interventions implemented to achieve this goal, but did not specify how those interventions delivered transitional care components to support the actual linkage of patients across studies [10]. Even studies of the widely-adopted substance use intervention Screening, Brief Intervention, and Referral to Treatment (SBIRT), rarely describe or evaluate practices used to refer or transition patients to ongoing care [11]. Such “transition strategies” (as we refer to from now on) are often vaguely described as linkages, discharge planning, care navigation, warm handoffs, or referrals to community care, but have never been characterized or compared in the context of hospital patients with SUD. This gap may be in part due to the challenge of comparing heterogenous interventions and practices that often use vague or inconsistent terminology. As such, more work is needed to explore the ‘black box’ of transition strategies used to support the ‘Cascade of Care’ [14] from the ED or inpatient setting to ongoing community-based SUD treatment (Figure 1).

**Figure 1:**
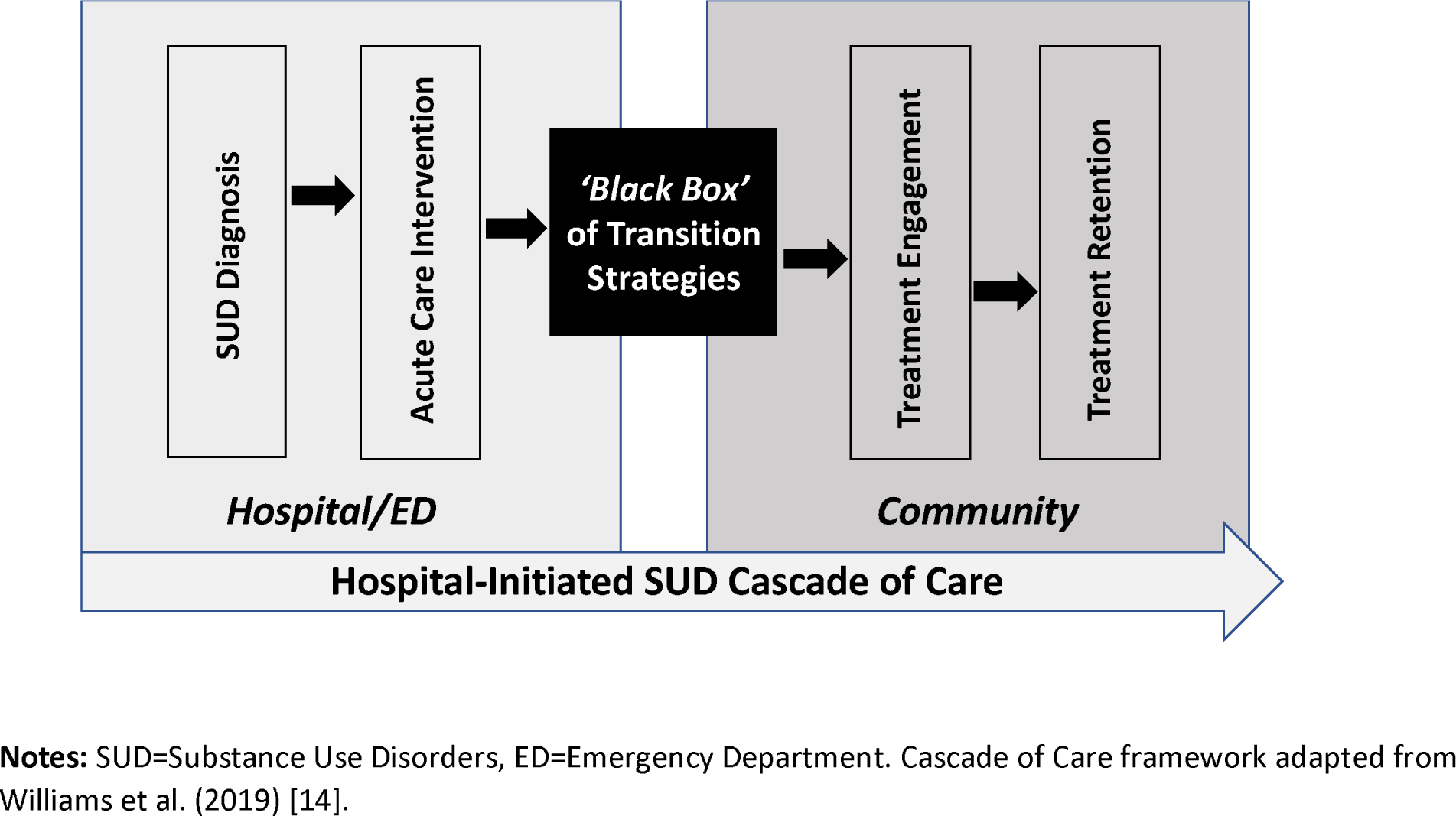
Conceptual Framework: The ‘Black Box’ of SUD Transition Strategies

Evidence on the most successful transition strategies can help health systems adopt the most effective and efficient practices to improve SUD outcomes among their patient populations. For OUD patients, specifically, improving transitions from hospital-to community-based provisions of care with evidence-based medication for OUD (MOUD) can significantly support patients’ success across the OUD Cascade of Care[15]. However, to do so, more research is needed to identify and categorize particular SUD care transition strategies, understand and break down their components, and identify the barriers and facilitators to their implementation and execution. To begin addressing this gap, the current scoping review aims to: 1) synthesize information from published literature on existing transition strategies used to link patients with SUDs from ED or inpatient hospital settings to community-based SUD treatment; 2) Group and classify transition strategies based on similar characteristics, forming an initial typology of strategies; 3) Summarize the primary barriers and facilitators encountered in implementing transition strategies within the studied programs and interventions.

## Methods

This scoping review was conducted in alignment with the Preferred Reporting Items of Systematic Reviews and Meta-Analyses for Scoping Review (PRISMA-ScR) checklist [16], as available in Supplemental Table 1.

### Search Strategy

An initial search strategy was developed using combinations of keywords related to SUDs and linkage to community-based SUD treatment from acute-care settings. The search strategy was further refined in consultation with an expert librarian at the New York University Health Sciences Library and adapted to the following databases: PubMed, EMBASE via OVID, CINAHL via EBSCO, and PsycInfo via OVID. The full search strategy for each database can be found in Supplemental Table 2. The final search was conducted on October 2, 2021. A manual search of the reference lists of included articles was conducted to capture potentially relevant studies that were not identified by the database search. Lastly, we reviewed the studies included in a recently published systematic review on acute-care interventions for SUD patients [10] to ensure a robust inclusion toward our aims.

### Eligibility Criteria

Articles were included if they (1) were published between January 1, 2000 and October 2, 2021, (2) described programs or interventions for adult patients with SUDs (excluding tobacco) hospitalized in general acute-care settings (i.e. either ED or inpatient hospital settings, not including specialized detox or psychiatric inpatient care), (3) evaluated post-hospital SUD treatment engagement outcomes, and (4) described practices for supporting patients’ transition from the acute-care setting to community SUD treatment. Given the review’s primary focus on characterizing strategies employed to support patients’ transition between acute-care to community SUD settings, the last inclusion criteria required the article to have a minimum of one sentence describing the transition strategy. Thus, otherwise eligible studies that failed to include a description of how patients were linked with post-discharge care were not included (one third of full text studies (n=51), Figure 2). While our search was not limited to English-language articles, the few non-English studies deemed eligible for full-text review and translated by team members, did not meet full inclusion criteria.

**Figure 2:**
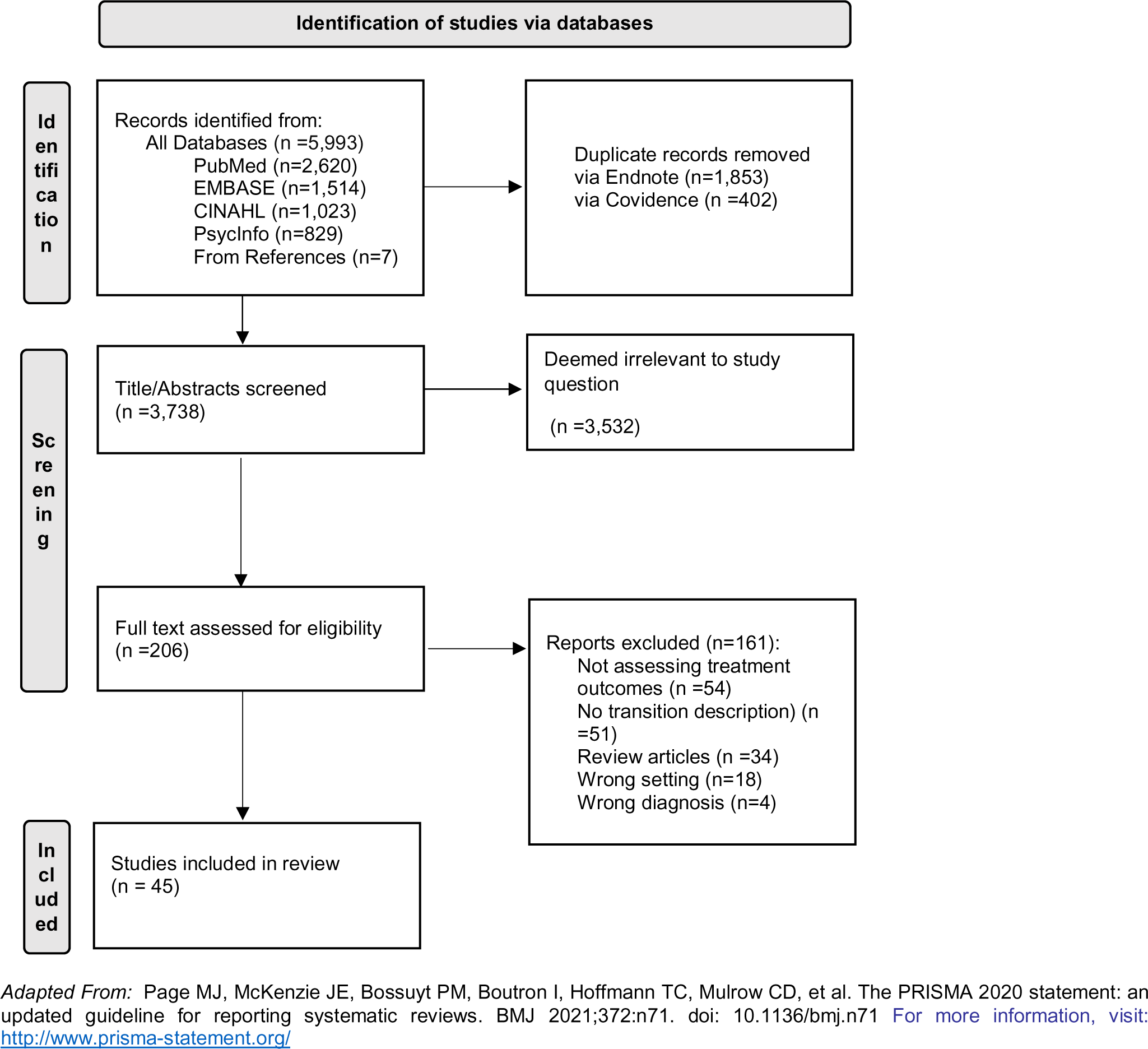
PRISMA diagram of study selection process

### Screening and Data Extraction

Database search results were first imported into Endnote for initial deduplication. The remaining records were uploaded into Covidence, an online subscription-based systematic review tool, where a second deduplication procedure was completed. All abstracts were screened by two independent reviewers from the study team with full blinding for initial eligibility. Disagreements were resolved through team discussions. Articles meeting initial eligibility were reviewed in full by four members of the team (BDR, MG, Y-HC, SN). Regular team discussions with the lead author (NK) facilitated final decisions on the inclusion of articles and their relevance to study aims.

A data extraction form was created and implemented through Covidence to systematically collect relevant information from included studies. The data extraction form was iteratively developed such that an initial form, based on study goals, was refined through multiple pilot rounds to extract relevant data from articles on transition strategies described across interventions until consensus was reached. Study information extracted from each article included author and month of publication; intervention setting (i.e., ED or inpatient); country; program/intervention name; target SUD population (e.g., OUD, AUD, etc.); study design and sample size; and evaluated post-discharge SUD treatment outcomes. For each article, detailed information was extracted on any described “transition strategies,” which we defined as practices (i.e. procedures or activities) undertaken to help link patients from the acute-care setting to community-based treatment at discharge. We also extracted information on other relevant intervention components described at the acute-care phase (location and types of staff delivering care in the acute-care setting, use of medications or brief interventions), the transition phase (location and types of staff involved in facilitating transition) and community treatment phase – including the SUD services or settings patients were being transitioned to (e.g. specialty SUD treatment programs, bridge clinics (generally low-barrier short-term programs to treat OUD while connecting patients to community resources[17])). Finally, we extracted information on any reported barriers or facilitators to support the transition of patients from acute-care to ongoing treatment in community-based settings.

### Synthesis of Intervention Components and Transition Strategies

We applied a narrative synthesis approach[18] to review the studies of acute-care interventions and the range of practices they employ to support the transition of patients to community-based SUD care. To do this, the lead author (NK) conducted an initial content analysis of each article focusing specifically on language related to transitioning patients from the acute-care to the community setting. The content was coded inductively and categorized into a typology of common “transition strategy’’ categories based on similar practices that emerged across the individual programs and interventions (e.g. scheduling appointment with a particular provider, providing transportation assistance to treatment appointment). Typology categorizations were discussed with the full team and iteratively modified based on team members’ expertise, until consensus was reached. To ensure consistency, a second member of the team (BR) double coded all transition strategies based on the final typology categories. The frequency of each transition strategy was then tallied across studies to characterize their distribution across the explored interventions in the ED and inpatient hospital settings.

### Synthesis of Primary Barriers and Facilitators across Transitions

Finally, to gain a better understanding of the implementation challenges involved in the transition of patients from acute-to community-based care, we conducted an analysis of the barriers and facilitators reported in the included articles pertaining specifically to the transition of patients across settings. Barriers and facilitators were organized based on five domains described in the Consolidated Framework for Implementation Research (CFIR)[19], including “intervention characteristics” (e.g. care coordination procedures, hospital-community provider partnerships), “inner setting” (resources for addiction care; provider time constraints), “outer setting” (regulation of MOUD; availability of MOUD providers), “individual characteristics” (provider attitudes; training in addiction care) and “implementation process” (leadership engagement; outcome evaluation).

## Results

A total of 3,738 unique records were identified across databases based on title and abstract screening. Of these, 206 were selected for full-text review, and 45 met full inclusion criteria. The PRISMA diagram outlining the study inclusion process is presented in Figure 2.

### Characteristics of Included Studies

An overview of characteristics of included studies is presented in Table 1, with details of each study available in Supplemental Table 3. Due to the distinct nature of ED and inpatient hospital interventions and settings, we present findings overall and by setting type. Of the 45 studies identified, 56% (n=25) described interventions taking place in the ED, 38% (n=17) in inpatient hospital settings, and 7% (n=3) interventions taking place in both ED and inpatient hospital settings. A large majority (n=38, 84%) of studies were U.S.-based. Study design varied, with the majority being observational studies (n=19, 42%) followed by pilot/feasibility studies (n=16, 36%) and randomized control trials (n=10, 22%). Interventions commonly targeted patients with OUD (n=32, 71%), followed by patients with AUD (n=15, 33%), and patients with SUD generally (n=14, 31%). Studies reported on multiple outcome measures related to linkage to community-based SUD treatment: The most common measures were whether there was any visit following discharge (n=28, 62%) and length of time retained in treatment post discharge (n=30, 67%) with some reporting time to first visit post-discharge (n=3, 7%). The exact definitions and metrics used to quantify the treatment engagement measures varied widely across articles, making intervention effects incomparable across studies. There was also great variability in the methods used to ascertain data on treatment engagement and in the reporting quality of these methods, which ranged from active data collection through contact with SUD providers (n=9, 20%), patients (n=6, 13%), or unspecified sources (n=14, 31%), to relying on existing data available through EHR (n=23, 51%) or other sources (n=6, 13%).

**Table 1:** Setting, target population and outcomes/study design by intervention setting

|  | **Overall n(%)**<br>**N=45 (100%)** | **ED n(%)**<br>**N=28 (62%)<sup>a</sup>** | **Hospital n(%)**<br>**N=20 (44%)<sup>a</sup>** |
| --- | --- | --- | --- |
| **Country** |  |  |  |
| United States | 38 (84.4) | 21 (75.0) | 15 (75.0) |
| Canada | 4 (8.9) | 3 (10.7) | 1 (5.0) |
| Other<sup>b</sup> | 3 (6.7) | 2 (7.1) | 1 (5.0) |
| **Study Design** |  |  |  |
| Randomized trial | 10 (22.2) | 4 (14.3) | 8 (40.0) |
| Observational study | 19 (42.2) | 14 (50.0) | 5 (25.0) |
| Pilot/feasibility study | 16 (35.6) | 10 (35.7) | 7 (35.0) |
| **Substance use Disorder/s Addressed<sup>c</sup>** |  |  |  |
| Substance use disorder (general) | 14 (31.1) | 10 (35.7) | 5 (25.0) |
| Opioid use disorder | 32 (71.1) | 22 (78.6) | 12 (60.0) |
| Alcohol use disorder | 15 (33.3) | 6 (21.4) | 10 (50.0) |
| Cocaine use disorder | 3 (6.7) | 0 (0.0) | 3 (15.0) |
| Methamphetamine use disorder | 2 (4.4) | 0 (0.0) | 2 (10.0) |
| Sedative use disorder | 2 (4.4) | 2 (7.1) | 0 (0.0) |
| **SUD Treatment Outcome/s Measured<sup>c</sup>** |  |  |  |
| Community SUD treatment visit post-discharge | 28 (62.2.0) | 23 (82.1) | 9 (45.0) |
| Time to first community SUD treatment visit | 3 (6.7) | 2 (7.1) | 1 (5.0) |
| Retention/length of engagement in SUD treatment | 30 (66.7) | 17 (60.7) | 14 (70.0) |
| **Data Sources Used to Ascertain Outcomes<sup>c</sup>** |  |  |  |
| *Active Data Collection* |  |  |  |
| Data collected from community treatment clinic | 9 (20.0) | 5 (17.9) | 4 (20.0) |
| Data collected via follow up up with patients | 6 (13.3) | 2 (7.1) | 4 (20.0) |
| Data collected via unspecified source | 14 (31.1) | 9 (32.1) | 8 (40.0) |
| *Passive Data Collection* |  |  |  |
| Data ascertained via Electronic Health Record | 23 (51.1) | 16 (57.1) | 7 (35.0) |
| Data ascertained via Prescription Drug Monitoring Programs | 2 (4.4) | 2 (7.1) | 0 (0.0) |
| Data ascertained via Medicaid claims | 1 (2.2) | 0 (0.0) | 1 (5.0) |
| Data ascertained via other administrative data<sup>d</sup> | 3 (6.7) | 3 (10.7) | 0 (0.0) |

Note: ED=Emergency Department, SUD=Substance Use Disorder,
<sup>a</sup> Three studies included patients from both the ED and inpatient setting
<sup>b</sup> Other countries include Switzerland (1), Spain (1), Denmark (1)
<sup>c</sup> Not mutually exclusive, will not add to 100%
<sup>d</sup> Other administrative data includes trauma registries, behavioral health databases

### Typology of Transition Strategies

The content analysis and inductive coding process resulted in a typology of 10 transition strategies that encompass practices to facilitate patients’ transition between acute-care and community-based SUD treatment settings. These were further divided into five “pre-discharge transition strategies”- which occur prior to patients leaving the acute-care setting - and five “post-discharge transition strategies” - which occur after the patient has already left the acute-care setting. Table 2 presents the typology of transition strategies, a description of each, and an example from the reviewed literature. Identified pre-discharge transition strategies included: Discuss Treatment Options; Schedule Appointment; Provider List; Electronic Referral; and General Linkage to Treatment. Identified post-discharge transition strategies included: Bridge Prescription; Transportation Assistance; Follow up Calls/Texts; Care Navigation; and Peer Support. Transition strategies described in each study were not mutually exclusive: In many cases, an intervention may have combined 2-3 of these strategies within the same intervention or tested multiple different strategies across intervention and control groups (see Supplemental Table 3 for the transition strategies identified in each study). For example, an interprofessional addiction consult service might discuss treatment options and schedule post-hospital appointments before discharge, and offer bridging medication prescription or peer support after discharge [12]. In addition, while categorized based on similarity of approach, the settings, staff involved, intensity of each transition strategy often varied substantially across studies, which could influence transition outcomes. For example, in some interventions, staff that supported care transitions such as peer navigators met patients in the hospital and continued working with them post-discharge [20], while in others, care navigators supporting transitions only contacted patients following discharge via phone calls[21]. In many cases, the level of detail on the actual transition practices involved were rather vague or unspecified and therefore difficult to extract and compare across interventions.

**Table 2:**
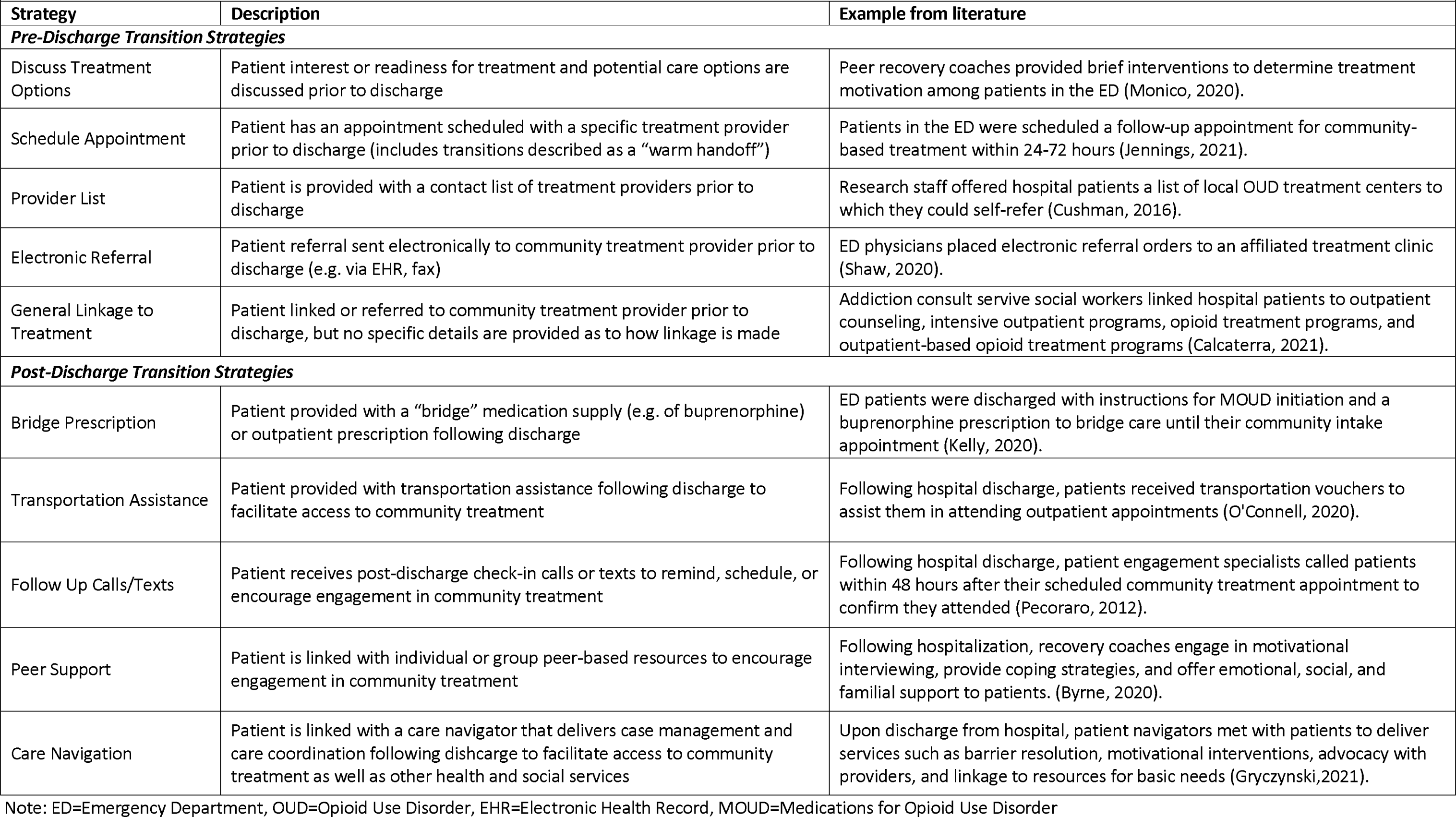
Typology of Transition Strategies Identified in the Literature

Figure 3 presents the frequency of pre-discharge and post-discharge strategies described across included interventions taking place in ED and inpatient hospital settings. For both ED and inpatient hospital settings, the most common pre-discharge transition strategy was scheduling an appointment, which was mentioned in 57% (n=16) of ED interventions and 50% (n=10) of inpatient interventions. In as many as 36% (n=10) of ED interventions and 30% (n=6) of inpatient interventions, there was a general mention of pre-discharge linkage of patients to SUD treatment programs or providers in the community, but there was no specific information on how this linkage was made or supported. While providing a bridge medication prescription was the most common post-discharge strategy mentioned in the ED setting (36%, n=10), it was much less commonly noted in the inpatient setting (20%, n=4. Care navigation and transportation assistance were more common in inpatient interventions (30%, n=6, for both) than ED settings (25%, n=7 and 18%, n=5, respectively). These differences may reflect the different circumstances and resources available across ED and inpatient settings, including the longer discharge planning period that is often available in inpatient settings and not ED settings.

**Figure 3:**
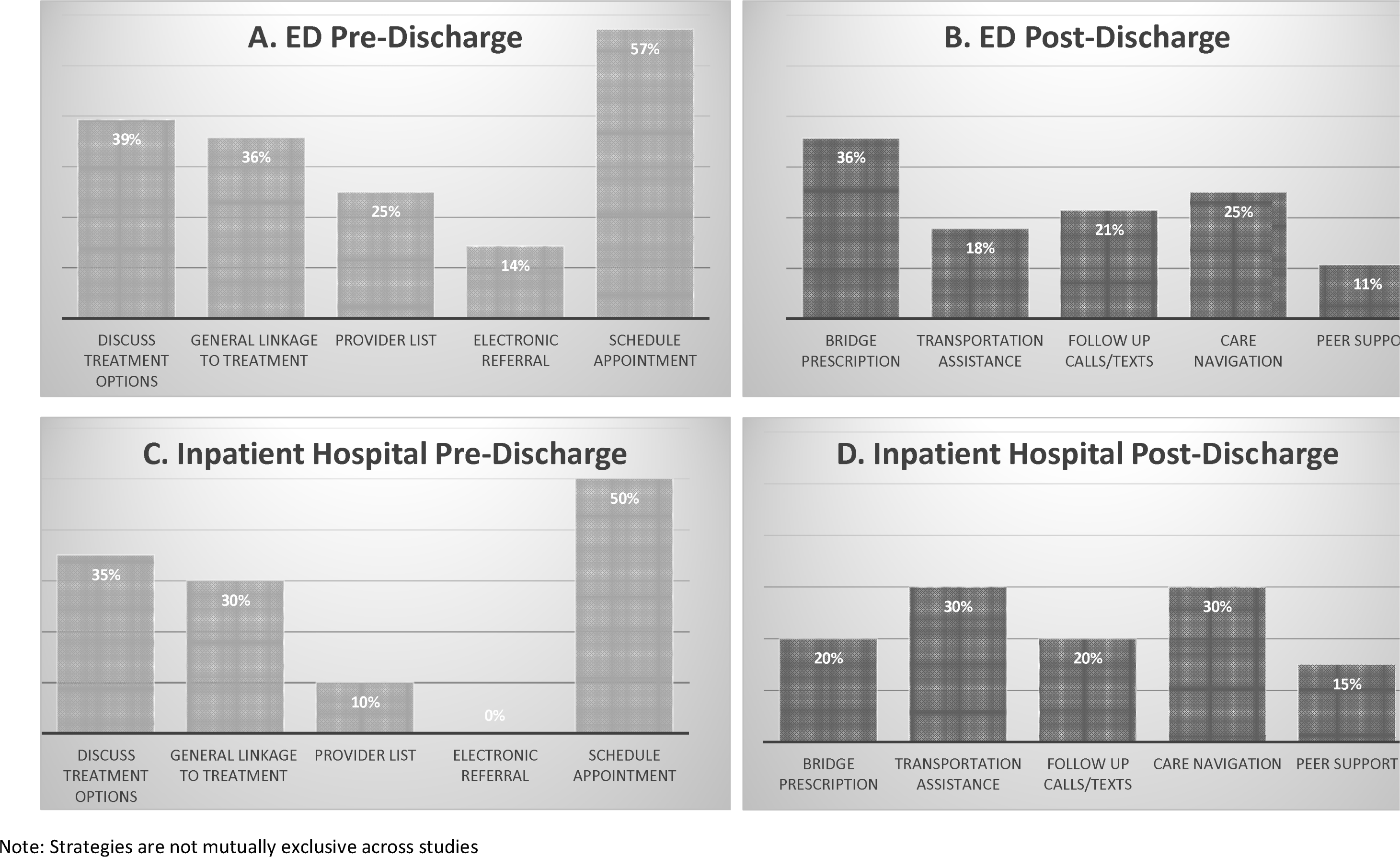
Frequency of strategies described across included interventions categorized as (A) pre-discharge strategies and (B) post-discharge strategies in ED interventio (n=28), and (C) pre-discharge and (D) post-discharge strategies in inpatient hospital interventions (n=20)

### Additional Intervention Components across Care Continuum

For each study, we identified and organized additional relevant components of the described interventions across the care continuum from the acute-care phase in the ED/hospital, to the transition phase supporting acute-care to community treatment, and the community treatment phase where patients are linked (Table 3).

**Table 3:** Components of intervention during acute care, transition, and community treatment phases

| Components of Intervention |  | Overall<br>N=45 (100%) | ED<br>N=28 (62%)<sup>a</sup> | Hospital<br>N=20 (44%)<sup>a</sup> |
| --- | --- | --- | --- | --- |
| Acute Care Phase | **Staff Involved in Acute Care Intervention<sup>b</sup>** |  |  |  |
|  | Multidisciplinary addiction care team | 11 (24.4) | 4 (14.3) | 9 (45.0) |
|  | Nurse | 10 (22.2) | 6 (21.4) | 3 (15.0) |
|  | Social worker/case manager | 10 (22.2) | 4 (14.3) | 7 (35.0) |
|  | Patient navigator | 5 (11.1) | 4 (14.3) | 1 (5.0) |
|  | Addiction counselor | 6 (13.3) | 4 (14.3) | 3 (15.0) |
|  | Peer | 3 (6.7) | 2 (7.1) | 2 (10.0) |
|  | Medical doctor | 22 (48.9) | 11 (39.3) | 12 (60.0) |
|  | Nurse practitioner or physician's assistant | 4 (8.9) | 2 (7.1) | 2 (10.0) |
|  | Other<sup>c</sup> | 3 (6.7) | 3 (10.7) | 2 (10.0) |
|  | Not specified | 11 (24.4) | 9 (32.1) | 1 (5.0) |
|  | **Location of Acute Care Staff** |  |  |  |
|  | Work directly in ED/Hospital unit | 34 (75.6) | 24 (85.7) | 12 (60.0) |
|  | Called upon as a part of a hospital consult team | 5 (11.1) | 1 (3.6) | 5 (25.0) |
|  | Not specified | 6 (13.3) | 3 (10.7) | 3 (15.0) |
|  | **Medication Initiation Offered<sup>b</sup>** |  |  |  |
|  | Buprenorphine | 26 (57.8) | 16 (57.1) | 11 (55.0) |
|  | Methadone | 10 (22.2) | 1 (3.6) | 10 (50.0) |
|  | Naltrexone | 6 (13.3) | 1 (3.6) | 5 (25.0) |
|  | Pharmacotherapy (unspecified) | 1 (2.2) | 0 (0.0) | 1 (5.0) |
|  | None | 14 (31.1) | 9 (32.1) | 6 (30.0) |
|  | **Brief Interventions Involved** |  |  |  |
|  | Screening, Brief Intervention, Referral to Treatment (SBIRT) | 7 (15.6) | 5 (17.9) | 2 (10.0) |
|  | Other Brief Intervention | 4 (8.9) | 4 (14.3) | 0 (0.0) |
| Transition Phase | **Staff Involved in Facilitating Transition<sup>b</sup>** |  |  |  |
|  | Multidisciplinary addiction care team | 4 (8.9) | 2 (7.1) | 2 (10.0) |
|  | Nurse | 11 (24.4) | 7 (25.0) | 4 (20.0) |
|  | Social Worker/case manager | 15 (33.3) | 6 (21.4) | 10 (50.0) |
|  | Patient navigator | 5 (11.1) | 3 (10.7) | 2 (10.0) |
|  | Addiction counselor | 5 (11.1) | 3 (10.7) | 5 (25.0) |
|  | Peer | 11 (24.4) | 6 (21.4) | 5 (25.0) |
|  | Medical doctor | 25 (55.6) | 18 (64.3) | 8 (40.0) |
|  | Nurse practitioner or physician's assistant | 3 (6.7) | 2 (7.1) | 1 (5.0) |
|  | Research assistant | 3 (6.7) | 3 (10.7) | 0 (0.0) |
|  | Other<sup>d</sup> | 4 (8.9) | 3 (10.7) | 2 (10.0) |
|  | Not specified | 4 (8.9) | 4 (14.3) | 0 (0.0) |
|  | **Location of Transition Staff** |  |  |  |
|  | Work directly in the ED/Hospital unit | 33 (73.3) | 23 (82.1) | 10 (50.0) |
|  | Employed by external organization/research team | 9 (20.0) | 4 (14.3) | 7 (35.0) |
|  | Called upon as a part of a hospital consult team | 4 (8.9) | 1 (3.6) | 3 (15.0) |
| Community Treatment Phase | **SUD Services Patients Transitioned To<sup>b</sup>** |  |  |  |
|  | SUD treatment providers (general) | 26 (57.8) | 12 (42.9) | 14 (70.0) |
|  | Treatment with MOUD | 26 (57.8) | 17 (60.7) | 11 (55.0) |
|  | Primary care provider | 5 (11.1) | 4 (14.3) | 1 (5.0) |
|  | Detoxification program | 4 (8.9) | 3 (10.7) | 3 (15.0) |
|  | Residential treatment program | 8 (17.8) | 4 (14.3) | 5 (25.0) |
|  | Bridge or other short-term clinic | 7 (15.6) | 4 (14.3) | 3 (15.0) |
|  | Peer support/self-help groups | 3 (6.7) | 1 (3.6) | 3 (15.0) |
|  | Other medical and mental health services<sup>f</sup> | 6 (13.3) | 5 (17.9) | 3 (15.0) |
|  | Harm reduction services | 2 (4.4) | 0 (0.0) | 2 (10.0) |
|  | Other community/social services<sup>g</sup> | 11 (24.4) | 6 (21.4) | 7 (35.0) |

Note: ED=Emergency Department, SUD=Substance Use Disorder, MOUD= Medications for Opioid Use Disorder
<sup>a</sup> Three studies included patients from both the ED and inpatient setting
<sup>b</sup> Not mutually exclusive, will not add to 100%
<sup>c</sup> Other staff involved in acute care (3) include pharmacist, clinical psychologist, and medical student
<sup>d</sup> Other staff involved in transition (4) include pharmacist, toxicologist, psychoeducator, staff from the outpatient clinic in which patients are referred
<sup>e</sup> Other medical and mental health services include care management, injury care, mental and behavioural health
<sup>f</sup> Other community/social services include transportation, legal support, educational support, familial support, domestic violence hotlines, basic needs support (housing, food, employment, insurance, clothing)

The most common staff types involved at the acute-care phase were medical doctors (n=22, 49%) and multidisciplinary addiction care teams (n=11, 24%), which were much more common in inpatient hospital settings (n=9, 45%) than ED settings (n=4, 14%). Many studies (n=11, 24%), particularly in the ED setting (n=9, 32%), did not specify what staff types were involved in delivering acute-care. In the majority of studies, described staff worked directly within the hospital unit (n=34, 76%), while in some they were called upon as part of an in-hospital consult service (n=5, 11%). When we assessed whether medications or behavioral brief interventions were delivered in the acute-care setting, the majority of studies involved buprenorphine initiation (n=26, 58%) with some mentioning methadone (n=10, 22%) or naltrexone (n=6, 13%) initiation. Nearly a third (n=14, 31%) of interventions did not mention any medications for SUD. Using an SBIRT model was reported in 16% (n=7) of studies and 9% (n=4) noted other brief interventions - these were primarily implemented in the ED (n=9, 32%) rather than inpatient hospital settings (10%).

For the transition phase, we found that for some interventions, transitions were facilitated by the same staff that delivered the acute-care portion of the intervention, while others had additional or unique staff delivering these practices. The most common staff types mentioned in this phase were medical doctors (n=25, 56%), especially in ED settings (n=18, 64%), followed by social workers or case managers (n=15, 33%), which were more common in inpatient hospital setting (n=10, 50%). In a few cases (n=3, 7%), staff facilitating the transition were noted to be part of a study research team rather than an acute-care clinical team. The location of transition staff varied across ED and inpatient interventions, with ED interventions more commonly relying on staff within the ED to facilitate the transition (n=23, 82%) while inpatient interventions often relied on staff called upon as part of a consult team (n=3, 15%) or employed by an external organization (n=7, 35%). Examples of consult or externally-employed staff facilitating transitions included medical toxicologists [22], resource specialists[23], patient engagement specialists [24] and peer specialists [21,25,26].

Finally, we assessed the nature of community treatment settings to which the studied interventions linked SUD patients following discharge. Studies most commonly described their intervention as generally linking patients to SUD treatment providers, but the exact nature or setting was unspecified (n=26, 58%). Many mentioned linking patients to treatment with MOUD specifically (n=26, 58%), especially in ED interventions (n=17, 61%). Other treatment settings included detoxification programs (n=4, 9%), residential treatment programs, (n=8, 18%), bridge clinics (n=7, 16%), and primary care providers (n=5, 11%). In addition to SUD treatment services, 7% (n=3) linked to peer support or self-help groups, 13% (n=6) to other medical or mental health services, 7% 24% (n=11) linked patients to other community or social services, which was more common amongst inpatient interventions (n=7, 35%). Only 4% (n=2) of interventions (all in the inpatient setting) mentioned linking patients to harm reduction services.

### Barriers and Facilitators to Transitions of Care

Authors of included studies discussed a range of barriers and facilitators that they found to be influential to the success of supporting the transition of patients from acute-care to ongoing SUD care in the community. Findings are organized based on CFIR domains as follows:

#### Intervention characteristics

Multiple studies noted barriers related to planning and executing the transition intervention itself, such as limited financial resources for program operations [27], difficulty of building partnerships between hospital and community-based SUD programs [22] and managing logistics and communication between hospital and community treatment teams to coordinate follow-up care for patients discharged from acute-care settings [28,29]. Facilitators discussed to alleviate some of these barriers included leveraging existing resources of community-based providers [27], having the same community-based providers also work in the acute-care setting to build trust and continuity across settings [27,30,31], and maintaining a close working relationship and open lines of communication between acute-care staff and community providers [32].

#### Inner Setting and Individual Characteristics

Many also discussed barriers and facilitators related to the acute-care setting of where the interventions took place and the individuals that delivered these interventions. Barriers described included limited staff capacity and slow uptake of novel protocols in the landscape of busy hospital units [33,34], implicit bias against the SUD population among acute-care providers [35], and frequent undertreatment of withdrawal in these settings [36]. Some facilitators for delivering care in these settings included conducting educational sessions for hospital staff to improve awareness of interventions and introduce concepts of harm reduction and trauma-informed care [27], and having the intervention delivered by non-medical staff such as social workers [28], patient navigators [20], individuals in recovery or peer recovery coaches [26,32] or external staff from community based programs who could dedicate more time to patients [31]. Studies also mentioned the importance of coordinating services across hospital care teams to improve efficiency [37]and the value of having supervisors who could help support intervention staff in coordinating patient care [20,32].

#### Outer Setting

Studies described multiple factors influencing the success of the transition that related to the outer setting of community services and resources. Barriers included limited availability and capacity of community SUD treatment providers such as limited hours of operation [38,39], frequent staff turnover [29], and limited availability in rural settings [40]. Many also mentioned the multiple vulnerabilities and social risk factors that SUD patients face, including employment instability, lack of housing, limited transportation, criminal justice involvement and other medical needs[23,30,40–43]. For digital interventions, barriers were mentioned around use of technology or distrust of sharing sensitive information [44]. Multiple studies also mentioned challenges around patient disinterest or motivation to continue treatment [26,33,43,44]. Finally, many discussed structural barriers such as federal regulations that limit availability of MOUD [38,40], the segregation of SUD treatment services from other medical and healthcare services[23] and limited financial coverage for SUD services among some populations [45]. Some examples of facilitators to address some of these barriers included providing bridge medications and services on days where community clinics are closed [46]and co-locating or working with community-based services near the acute-care setting to facilitate access [47,48]. To address patient barriers, some discussed the importance of allowing for patient preference in choosing care options [35], creating service models that integrate SUD care with other medical, mental health and social supports [23] and dedicating monetary funds as part of care navigation support to address social challenges faced by patients leaving the hospital [20]. To address financial and structural barriers, some mentioned working with services where patients’ ability to pay did not influence service access [29]and pointed to the more flexible treatment policies and universal healthcare system in Canada that alleviated access and financial barriers relative to the US [49].

#### Implementation Process

While few explicitly discussed barriers and facilitators related to the process of implementation, one study did note that leadership involvement and early buy-in was a critical success factor during the implementation and post-implementation processes of their transition intervention [32].

## Discussion

The field of addiction medicine is at a critical time in which hospitals and health systems are paying greater attention to the care of patients with SUD. Growing research about the efficacy of acute-care interventions[6,7,50,51] along with advocacy on behalf of medical societies and other groups for health systems to improve care for SUD [8,52,53]have instigated a rapid expansion of these programs. Indeed, implementing acute-care interventions for overdose and SUD is noted in national recommendations to address the overdose crisis[54,55] and often a priority for resource allocation in the wake of the opioid lawsuit settlement funds[56,57]. Despite these advances, there is still much that remains unknown about what makes these hospital interventions and their implementation most effective. Specifically, while linkage to ongoing community treatment for SUD after hospitalization is almost always highlighted as a desired outcome, how this transition is best supported by these interventions is much less clear. As such, our paper aimed to begin addressing this gap by establishing a typology of existing strategies for linking SUD patients from acute-care settings to community-based treatment.

As expected, our study identified significant heterogeneity in practices for transitioning patients from the acute-care setting to community treatment, in both the procedures and activities (“transition strategies”) employed, as well as other components of interventions used to support these strategies, such as the staff types involved or setting to which patients were being transitioned. Moreover, across interventions, even similar practices were often delivered by staff with very different roles (e.g. medical doctor vs. social worker) or via different staffing models (e.g. consult service vs. in-unit staff). In some cases, interventions were delivered by external research staff, which raises questions about sustainability of these practices. There was also significant variability in the dose, intensity, and resources available for the transition strategies employed. Still, we were able to organize strategies into an initial typology based on recurrent approaches amongst the described interventions, which can serve as a basis for future work on the relative effectiveness of these strategies and how they might interact with other intervention components. Importantly, our study identified a distinction between transition strategies that occur prior vs. following patient discharge from the acute-care setting, which may have different implications for implementation and resource allocation.

An important finding of this scoping review is that it highlights the rather scarce information available in the scientific literature about the nature of transition strategies that occur as part of acute-care interventions for SUD. As many as one third of studies reviewed at the full-text phase (n=51) were excluded for not having a description of practices involved in transitioning patients from the hospital to community treatment. This is despite many still having community SUD treatment linkage as a primary outcome of their studied intervention. Even among studies that did include a description of the transition strategies, details of the specific activities or procedures, duration, or individuals involved in executing the transition strategy were often left out. One of the most common pre-discharge transition strategies identified in our typology in both ED and inpatient hospital interventions was “General Linkage to Treatment,” in which there was some type of linkage or referral involved, but there were no further details provided around how such linkage was facilitated. Even studies describing conducting a “warm-handoff”, often did not explain what this entailed. It is possible some of these details may have been previously published in program descriptions excluded from our review[12,58]. Still, this lack of detail reveals that the transition to community treatment itself has been underestimated in importance in research and potentially in intervention planning itself. This has been recognized in the past with SBIRT studies where the “RT-referral to treatment” portion of the intervention is rarely included or described[11]. And while much of the care coordination and care transitions literature even outside of SUD care is complex and somewhat opaque [59], the SUD transitions literature seems to be particularly obscure. These findings emphasize the need for the addiction medicine field to place greater emphasis on the transition portion of these interventions, a feat that likely requires greater collaboration across inpatient and outpatient care services.

Across reviewed studies, many implementation barriers and facilitators to supporting patient transitions were discussed, ranging from organizational characteristics specific to acute-care settings, to wider issues around provider availability and treatment policies that influence care options in the community. While hospital intervention teams did not always have power to eliminate these barriers, many described creative solutions, such as forming partnerships and workflows with community programs and trying to match needs and preferences of patients with services offered. Multiple prior studies have assessed barriers and facilitators to the implementation of ED and inpatient hospital-based interventions for SUD[60], but less attention has focused explicitly on the process of transition to ongoing community treatment[61,62]. Thus, more work is needed to understand and address barriers as they relate to the range of transition strategies identified in this review, how they are supported and reimbursed, and how they play out differently in ED vs. inpatient hospital settings. This understanding can help better guide resource allocation towards closing gaps in continuity of care, especially as hospital systems begin to integrate OUD Cascade of Care models and other mechanisms to track patient continuity across care settings[63,64] Critical to these efforts will be continuing to address existing barriers to retention in treatment once patients are linked with community-based care, which are frequently experienced among patients referred to treatment from ED or inpatient hospital settings [65–67].

It is important to place this study in the context of recent efforts to better streamline and characterize acute-care interventions for SUD, whose wide range of outcomes have been recently synthesized in a systematic review by James et al. (2023)[10]. In the inpatient setting, this has included efforts to compare and contrast different models of hospital-based care for SUD patients that range from interprofessional addiction consult models to community in-reach models, and vary based on target population, staff roles, clinical activities (including discharge planning), and larger systems change activities[7]. In the ED setting, multiple models of care have been described and compared, and often involve various combinations of MOUD prescribing, peer support or brief interventions, and referrals and bridge clinics to support patients’ continuity of care [68,69]. Further work is needed to characterize how these different models of care make use of particular transition strategies identified in this review, and how these strategies interact with other characteristics of acute-care interventions to successfully engage patients in ongoing care. While it may not be possible to disentangle all the individual components of these complex interventions to study their independent effects, thoughtfully characterizing these elements when designing and comparing interventions or models of care is critical for informing their effective implementation and sustainability. As such, the current review can set the stage for studying how certain transition strategies interact with other components of interventions and service models to support the transition of patients to community treatment. Finally, it is important to note that few interventions noted referring patients to harm reduction or other medical or mental health services. While this may have not been the primary goals of the included interventions, it is crucial that acute-care interventions for SUD patients, many whom may not be interested in treatment, better integrate harm reduction and other services to address risks and complex health needs of SUD patients[70].

Our study is subject to multiple limitations. First, our typology represents an attempt to classify SUD transition strategies based on peer-reviewed articles that typically describe only limited components of complex and heterogeneous interventions. The typology is therefore by nature reductive and only based on practices reported in the peer-reviewed literature, with the goal of forming an initial categorization scheme that would be more suitable for comparison and evaluation across multiple settings and programs. Our typology also does not take into account variation in dose, intensity, or resources allocated for the strategies described, which should be an important focus of future research. Second, as we limited our inclusion to studies that reported linkage outcomes, we may have missed articles that describe the intervention and the transition strategies in more detail, but were not captured in included articles. Thus, the transition strategies described here should be used to exemplify the breadth and types of strategies described in the literature, not provide a comprehensive list of possible transition strategies. Third, our study inclusion period covered a time period of over 20 years, during which there were many changes in the burden and need for SUD interventions, and in the clinical approaches to addressing SUDs during this time. This is especially notable in the rapid proliferation of overdose and OUD interventions involving MOUD over the last five years. Thus, while some of the older studies can still provide relevant insight into transition strategies that have been implemented in the past, some may not be as relevant to the most up-to-date clinical practice today. Finally, as our review was limited to peer-reviewed studies published before October of 2022, and as this is a rapidly evolving field, it is possible we may have missed other studies that were published more recently or that are available in the gray literature.

In sum, this scoping review aimed to review the literature and draw attention to SUD care transition strategies as a critical element of effective acute-care interventions. It is the hope that this initial typology and framework can help us begin to better describe and compare strategies implemented, toward more robust evaluations of programs and translation of effective interventions across care settings. Finally, improving our knowledge of care transition strategies is not only important in the context of acute-care interventions, but can serve to improve transitions of care among SUD patients across multiple touchpoints. Thus, future work should focus on how transition strategies may best support care continuity in SUD service utilization at other high-risk moments, such as at discharge from criminal legal settings or when transitioning from bridge clinics to long term primary care or other community-based care settings.

## Supporting information

Supplement_updated

## Data Availability

All data on studies included in this scoping review are publicly available via scientific databases

## Abbreviations

SUD: Substance use disorder
OUD: Opioid use disorder
MOUD: Medications for opioid use disorder
ED: Emergency department
CFIR: Consolidated framework for implementation research

## Declarations

### Ethics approval and consent to participate

Not applicable.

### Consent for publication

Not applicable.

### Availability of data and materials

Not applicable.

### Authors’ contributions

NK conceptualized the research study, conducted analysis of transition strategies and drafted the manuscript. BDR and MG led the database search, extracted data, drafted findings and edited the manuscript. YHC and SN screened studies for inclusion, extracted data, and edited the manuscript. JC, HE, and JM helped conceptualize the study, informed and contributed to analysis of transition strategies, and edited the manuscript.

## Acknowledgements

Authors thank Timothy Roberts from the NYU Health Sciences Library for his assistance in forming the initial search strategy for the scoping review.

