## Supplement_updated for "Strategies to support substance use disorder care transitions from acute-care to community-based settings: A Scoping review and typology"

**Supplemental Table 1: Preferred Reporting Items of Systematic Reviews and Meta-Analyses for Scoping Review (PRISMA-ScR) checklist**

| **SECTION** | **ITEM** | **PRISMA-ScR CHECKLIST ITEM** | **REPORTED ON PAGE #** |
| --- | --- | --- | --- |
| **TITLE** | | | |
| Title | 1 | Identify the report as a scoping review. | 1 |
| **ABSTRACT** | | | |
| Structured summary | 2 | Provide a structured summary that includes (as applicable): background, objectives, eligibility criteria, sources of evidence, charting methods, results, and conclusions that relate to the review questions and objectives. | 2 |
| **INTRODUCTION** | | | |
| Rationale | 3 | Describe the rationale for the review in the context of what is already known. Explain why the review questions/objectives lend themselves to a scoping review approach. | 4 |
| Objectives | 4 | Provide an explicit statement of the questions and objectives being addressed with reference to their key elements (e.g., population or participants, concepts, and context) or other relevant key elements used to conceptualize the review questions and/or objectives. | 5 |
| **METHODS** | | | |
| Protocol and registration | 5 | Indicate whether a review protocol exists; state if and where it can be accessed (e.g., a Web address); and if available, provide registration information, including the registration number. | n/a |
| Eligibility criteria | 6 | Specify characteristics of the sources of evidence used as eligibility criteria (e.g., years considered, language, and publication status), and provide a rationale. | 6 |
| Information sources* | 7 | Describe all information sources in the search (e.g., databases with dates of coverage and contact with authors to identify additional sources), as well as the date the most recent search was executed. | 5 |
| Search | 8 | Present the full electronic search strategy for at least 1 database, including any limits used, such that it could be repeated. | Appendix Table 2 |
| Selection of sources of evidence† | 9 | State the process for selecting sources of evidence (i.e., screening and eligibility) included in the scoping review. | 6 |
| Data charting process‡ | 10 | Describe the methods of charting data from the included sources of evidence (e.g., calibrated forms or forms that have been tested by the team before their use, and whether data charting was done independently or in duplicate) and any processes for obtaining and confirming data from investigators. | 6-7 |
| Data items | 11 | List and define all variables for which data were sought and any assumptions and simplifications made. | 7 |
| Critical appraisal of individual sources of evidence§ | 12 | If done, provide a rationale for conducting a critical appraisal of included sources of evidence; describe the methods used and how this information was used in any data synthesis (if appropriate). | n/a |
| Synthesis of results | 13 | Describe the methods of handling and summarizing the data that were charted. | 7-8 |
| **RESULTS** | | | |
| Selection of sources of evidence | 14 | Give numbers of sources of evidence screened, assessed for eligibility, and included in the review, with reasons for exclusions at each stage, ideally using a flow diagram. | 8, Figure 2 |
| Characteristics of sources of evidence | 15 | For each source of evidence, present characteristics for which data were charted and provide the citations. | 8-9 |
| Critical appraisal within sources of evidence | 16 | If done, present data on critical appraisal of included sources of evidence (see item 12). | n/a |
| Results of individual sources of evidence | 17 | For each included source of evidence, present the relevant data that were charted that relate to the review questions and objectives. | Appendix Table 3 |
| Synthesis of results | 18 | Summarize and/or present the charting results as they relate to the review questions and objectives. | 9-11 |
| **DISCUSSION** | | | |
| Summary of evidence | 19 | Summarize the main results (including an overview of concepts, themes, and types of evidence available), link to the review questions and objectives, and consider the relevance to key groups. | 13-16 |
| Limitations | 20 | Discuss the limitations of the scoping review process. | 16-17 |
| Conclusions | 21 | Provide a general interpretation of the results with respect to the review questions and objectives, as well as potential implications and/or next steps. | 17 |
| **FUNDING** | | | |
| Funding | 22 | Describe sources of funding for the included sources of evidence, as well as sources of funding for the scoping review. Describe the role of the funders of the scoping review. | 27 |

**Supplemental Table 2. Search Strategy**

| **Database** | **Strategy** |
| --- | --- |
| PubMed | ((Alcohol-Related Disorders/diagnosis[Mesh] OR Alcohol-Related Disorders/drug therapy[Mesh] OR Alcohol-Related Disorders/epidemiology[Mesh] OR Alcohol-Related Disorders/nursing[Mesh] OR Alcohol-Related Disorders/organization and administration[Mesh] OR Alcohol-Related Disorders/pharmacology[Mesh] OR  Alcohol-Related Disorders/rehabilitation[Mesh] OR  Alcohol-Related Disorders/therapy[Mesh] OR Alcohol-Related Disorders/prevention and control[Mesh] OR Alcoholism[tiab] OR "alcohol Abuse*"[tiab] OR  Substance-Related Disorders/diagnosis[Mesh] OR Substance-Related Disorders/drug therapy[Mesh] OR Substance-Related Disorders/epidemiology[Mesh] OR Substance-Related Disorders/nursing[Mesh] OR Substance-Related Disorders/pharmacology[Mesh] OR Substance-Related Disorders/organization and administration[Mesh] OR Substance-Related Disorders/prevention and control[Mesh] OR Substance-Related Disorders/rehabilitation[Mesh] OR Substance-Related Disorders/standards[Mesh] OR Substance-Related Disorders/therapy[Mesh] OR "SUD*" [tiab] OR "substance related disorder*"[tiab] OR Drug Overdose/drug therapy[Mesh] OR Drug Overdose/epidemiology[Mesh] OR Drug Overdose/nursing[Mesh] OR Drug Overdose/organization and administration[Mesh] OR Drug Overdose/prevention and control[Mesh] OR Drug Overdose/rehabilitation[Mesh] OR Drug Overdose/therapy[Mesh] OR Drug Overdose/diagnosis[Mesh] OR "Drug Overdose*"[tiab] OR "People who use drugs"[tiab] OR "Opioid Use Disorder"[tiab]) **AND** (Patient Discharge/methods[Mesh] OR Patient Discharge/organization and administration[Mesh] OR Patient Discharge/standards[Mesh] OR "Patient discharge*"[tiab] OR Patient Navigation/methods[Mesh] OR Patient Navigation/organization and administration[Mesh] OR Patient Navigation/standards[Mesh] OR Patient Navigation/methods[Mesh] OR "Patient navigat*"[tiab] OR Patient Care Planning/methods[Mesh] OR Patient Care Planning/organization and administration[Mesh] OR Patient Care Planning/standards[Mesh] OR "Patient Care Plan*"[tiab] OR Referral and Consultation/methods[Mesh] OR Referral and Consultation/organization and administration[Mesh] OR Referral and Consultation/standards[Mesh] OR “Referral”[tiab] OR "Consult*"[tiab] OR Continuity of Patient Care/methods[Mesh] OR Continuity of Patient Care/nursing[Mesh] OR  Continuity of Patient Care/organization and administration[Mesh] OR Continuity of Patient Care/pharmacology[Mesh] OR Continuity of Patient Care/standards[Mesh] OR Continuity of Patient Care/supply and distribution[Mesh] OR Continuity of Patient Care/therapy[Mesh] OR "Continuity of Patient Care"[tiab] OR ("Transition*"[tiab] AND ("Care"[tiab] OR "treatment*"[tiab])) OR ("Linkage*"[tiab] AND ("Care"[tiab] OR "Treatment*"[tiab])) OR ("Transfer*"[tiab] AND ("Care"[tiab] OR "Treatment*"[tiab])) OR ("Care"[tiab] AND ("Follow up"[tiab])) OR ("Cascade"[tiab] AND ("Care"[tiab] OR "Treatment*"[tiab])) OR ("Coordination"[tiab] AND ("Care"[tiab] OR "Treatment*"[tiab])) OR "Warm handoff"[tiab] OR "Bridge clinic"[tiab] OR "Addiction consult*"[tiab] OR "Addiction consult team*"[tiab] OR "Health care transition model"[tiab] OR "Consultation team"[tiab] OR "post discharge"[tiab] OR ("Post discharge"[tiab] AND ("navigator*"[tiab] OR "Care"[tiab] OR "Intervention*"[tiab]))) **AND** ("hospitals"[MeSH Terms] OR "hospital*"[tiab] OR "emergency service, hospital"[MeSH Terms] OR "emergency service*"[tiab] OR "emergency medical services"[MeSH Terms] OR "hospitalization"[MeSH Terms] OR "Acute care"[tiab] OR "detox*"[tiab]) **AND** (2000:2021[pdat])) |
| EMBASE via OVID | (exp alcoholism/di, dt, ep, pc, rh, th OR exp drug overdose/di, dt, ep, pc, rh, th OR exp "substance use"/th  OR (Alcoholism or "alcohol Abuse*" or "SUD*" or "substance related disorder*" or "Drug Overdose*" or "People who use drugs" or "Opioid Use Disorder").ti,ab.) **AND** (exp hospital discharge/ OR exp patient care planning/ OR exp patient referral/ OR exp patient care/ae OR ("Warm handoff" or "Bridge clinic" or "Addiction consult*" or "Addiction consult team*" or "Health care transition model" or "Consultation team" or "post discharge" or (Post discharge adj3 (navigator* or Care)) or ((Transition* adj3 (Care or treatment*)) or (Linkage* adj3 (Care or Treatment*)) or ("Transfer*" adj3 (care or Treatment)) or (Care adj3 Follow up) or (Cascade adj3 (Care or Treatment*)) or (Coordination adj3 (Care or Treatment))) or ("Patient discharge*" or "Patient navigat*" or "Patient Care Plan*" or "Continuity of Patient Care" OR "Referral" OR "Consult*")).ti,ab.) **AND** (exp hospital/ OR exp hospital emergency service/ OR exp emergency health service/ OR exp hospitalization/ OR ("hospital*" or "emergency service*" or "Acute care" or "detox*").ti,ab.) |
| CINAHL via EBSCO | ((MH "Alcohol-Related Disorders/DI/DT/EP/NU/TH/RH/PC/OG/ST") OR Alcoholism OR "alcohol Abuse*" OR (MH "Substance Use Disorders/DI/DT/EP/NU/RH/TH") OR "SUD*" OR "substance related disorder*" OR (MH "Overdose/DI/DT/EP/NU/OG/PC/RH/TH") OR "Drug Overdose*" OR "People who use drugs" OR "Opioid Use Disorder")) **AND** ((MH "Patient Discharge/MT/OG/ST") OR "Patient discharge*" OR (MH "Patient Navigation/MT/ST") OR "Patient navigat*" OR (MH "Patient Care Plans/MT/ST") OR "Patient Care Plan*" OR (MH "Referral and Consultation/MT/ST") OR "Referral" OR "Consult*" OR "Continuity of Patient Care" OR ("Transition*" AND ("Care" OR "treatment*")) OR ("Linkage*" AND ("Care" OR "Treatment*")) OR ("Transfer*" AND ("Care" OR "Treatment*")) OR ("Care" AND ("Follow up")) OR ("Cascade" AND ("Care" OR "Treatment*")) OR ("Coordination" AND ("Care" OR "Treatment*")) OR "Warm handoff" OR "Bridge clinic" OR "Addiction consult*" OR "Addiction consult team*" OR "Health care transition model" OR "Consultation team" OR "post discharge" OR ("Post discharge" AND ("navigator*" OR "Care" OR "Intervention*"))) **AND (**(MH "Hospitals") OR "hospital*" OR (MH "Emergency Service") OR "emergency service*" OR (MH "Emergency Medical Services") OR (MH "Hospitalization") OR "Acute care" OR "detox*") |
| PsycINFO via OVID | (exp Alcoholism/ OR exp "Substance Use Disorder"/ OR exp Drug Overdoses/ OR (Alcoholism or "alcohol Abuse*" or "SUD*" or "substance related disorder*" or "Drug Overdose*" or "People who use drugs" or "Opioid Use Disorder").ti,ab.) **AND** (exp Hospital Discharge/ OR exp Treatment Planning/ OR exp Professional Referral/ OR exp "Continuum of Care"/ OR ("Warm handoff" or "Bridge clinic" or "Addiction consult*" or "Addiction consult team*" or "Health care transition model" or "Consultation team" or "post discharge" or (Post discharge adj3 (navigator* or Care)) or ((Transition* adj3 (Care or treatment*)) or (Linkage* adj3 (Care or Treatment*)) or ("Transfer*" adj3 (care or Treatment)) or (Care adj3 Follow up) or (Cascade adj3 (Care or Treatment*)) or (Coordination adj3 (Care or Treatment))) or ("Patient discharge*" or "Patient navigat*" or "Patient Care Plan*" or "Continuity of Patient Care" or "Referral" or "Consult*")).ti,ab.) **AND** (exp Hospitals/ OR exp Emergency Services/ OR exp Hospitalization/ OR ("hospital*" or "emergency service*" or "Acute care" or "detox*").ti,ab.) |

**Supplemental Table 3. Summary Table**

| **Author, Year** | **Transition Strategy** | **Intervention Setting** | **Primary SUD Addressed** | **Brief Behavioral Intervention in Hospital** | **Medications Initiated in Hospital** | **Staff Before Transition** | | **Staff During Transition** | | **SUD Services Patients Transitioned To** |
| --- | --- | --- | --- | --- | --- | --- | --- | --- | --- | --- |
|  |  |  |  |  |  | **Staff Involved** | **Staff Location** | **Staff Involved** | **Staff Location** |  |
| Anderson, 2021 [1] | Pre-discharge discuss treatment options; Post-discharge care navigation | ED | AUD | Yes | Naltrexone | Medical doctor; Patient navigator | Work directly in ED/Hospital | Patient navigator | Placed directly in ED/hospital | Bridge/short term clinic |
| Beauchamp, 2021 [2] | Post discharge bridge prescription ; Post-discharge transportation assistance; Post-discharge care navigation; Post discharge peer support | ED; Inpatient hospital | SUD (general); OUD | No | Buprenorphine | Nurse; Social worker/case manager; Multidisciplinary addiction care team; Medical doctor | Work directly in ED/Hospital | Addiction counselor; Medical doctor; Toxicologists | Employed by external organization | Treatment with MOUD; Detox program; Residential/inpatient treatment providers; Other medical and mental health services; Other community resources |
| Beieler, 2021 [3] | General linkage to treatment;Post discharge bridge prescription | Inpatient hospital | OUD | No | Methadone; Buprenorphine | Multidisciplinary addiction care team; Addiction counselor | Not specified | Social worker/case manager; Multidisciplinary addiction care team | Called upon to ED/hospital as a consult | Treatment with MOUD; Other medical and mental health services; Other community resources |
| Berger, 2017 [4] | Pre-discharge discuss treatment options; Pre-discharge schedule appointment; Post-discharge transportation assistance; Post-discharge care navigation | Inpatient hospital | AUD | Yes | None | Addiction counselor | Called upon as part of Addiction consult team | Social worker/case manager; Addiction counselor | Employed by external organization | SUD treatment providers (unspecified) |
| Blanchette-Martin, 2016 [5] | General linkage to treatment | ED | SUD (general); AUD | No | None | Nurse; Social worker/case manager; Medical doctor | Work directly in ED/Hospital | Nurse; Social worker/case manager; Psychoeducator | Placed directly in ED/hospital | SUD treatment providers (unspecified); Residential/inpatient treatment providers |
| Bogan, 2020 [6] | Pre-discharge discuss treatment options; General linkage to treatment | ED | SUD (general); OUD | Yes | Buprenorphine | Patient navigator; Nurse; Medical doctor | Work directly in ED/Hospital | Nurse; Patient navigator; Peer; Medical doctor | Placed directly in ED/hospital | SUD treatment providers (unspecified); Treatment with MOUD |
| Brothers, 2021 [7] | General linkage to treatment, Post discharge bridge prescription | Inpatient hospital | OUD | No | Methadone; Buprenorphine | Medical doctor | Work directly in ED/Hospital | Medical doctor | Called upon to ED/hospital as a consult | SUD treatment providers (unspecified); Treatment with MOUD; Harm reduction services; Other community resources |
| Byrne, 2020 [8] | Post discharge peer support; Post-discharge transportation assistance; Post-discharge follow up calls; Post-discharge care navigation | Inpatient hospital | SUD (general); AUD | No | None | Social worker/case manager; Medical doctor; Medical student | Work directly in ED/Hospital | Peer; Medical doctor | Employed by external organization | SUD treatment providers (unspecified); Other community resources |
| Calcater­ra, 2021 [9] | Pre-discharge discuss treatment options; General linkage to treatment | Inpatient hospital | OUD; AUD; Methamphetamine & other stimulant use disorder | No | Methadone; Buprenorphine; Naltrexone | Social worker/case manager; Medical doctor | Called upon as part of Addiction consult team | Social worker/case manager | Work directly in ED/Hospital | SUD treatment providers (unspecified); Treatment with MOUD; Residential/inpatient treatment providers |
| Cushman, 2016 [10] | Pre-discharge schedule appointment; Pre-discharge provider list | Inpatient hospital | OUD | No | Buprenorphine | None specified | Work directly in ED/Hospital | Nurse; Medical doctor | Work directly in ED/Hospital | Treatment with MOUD |
| D'Onofrio, 2010 [11] | Pre-discharge discuss treatment options Post-discharge transportation assistance; Pre-discharge schedule appointment; General linkage to treatment | ED | SUD (general); AUD | Yes | None | Patient navigator | Work directly in ED/Hospital | Patient navigator | Work directly in ED/Hospital | SUD treatment providers (unspecified); Primary care providers; Other community resources |
| D'Onofrio, 2015 [12] | Pre-discharge provider list; Pre-discharge discuss treatment options; Pre-discharge schedule appointment; Post-discharge transportation assistance; Post discharge bridge prescription | ED | OUD | Yes | Buprenorphine | Medical doctor; Research Associate | Work directly in ED/Hospital | Medical doctor; Research assistant | Work directly in ED/Hospital | Treatment with MOUD |
| D'Onofrio, 2017 [13] | Pre-discharge provider list; Post discharge bridge prescription; Pre-discharge discuss treatment options; Pre-discharge schedule appointment; Post-discharge transportation assistance ; | ED | OUD | Yes | Buprenorphine | Medical doctor; Research Assistant | Work directly in ED/Hospital | Medical doctor; Research Associate | Work directly in ED/Hospital | SUD treatment providers (unspecified); Treatment with MOUD; Primary care providers; Detox program; Residential/inpatient treatment providers |
| Dahlem, 2020 [14] | Post-discharge follow up calls; Post-discharge care navigation; Post-discharge peer support; | ED | OUD | No | None | Social worker/case manager; Patient navigator | Not specified | Social worker/case manager; Patient navigator; Peer | Employed by external organization | Treatment with MOUD; Detox program; Other medical and mental health services; Other community resources |
| Dunkley, 2019 [15] | Pre-discharge discuss treatment options; Post discharge bridge prescription ; Pre-discharge schedule appointment | ED | OUD | No | Buprenorphine | Addiction counselor; Medical doctor | Work directly in ED/Hospital | Nurse; Medical doctor; Nurse practitioner; Physician assistant | Work directly in ED/Hospital | Treatment with MOUD |
| Edwards, 2020 [16] | Pre-discharge schedule appointment; General linkage to treatment | ED | OUD | No | Buprenorphine | None specified | Work directly in ED/Hospital | Nurse; Medical doctor | Work directly in ED/Hospital | Treatment with MOUD |
| Englander, 2019 [17] | Pre-discharge schedule appointment | Inpatient hospital | SUD (general) | No | Buprenorphine; Methadone; Naltrexone | Social worker; coordinator; Medical doctor; peer | Work directly in ED/Hospital | Social worker; Peer; Medical doctor | Work directly in ED/Hospital | SUD treatment providers (unspecified) |
| Gryczynski, 2021 [18] | General linkage to treatment;Pre-discharge discuss treatment options; Post-discharge care navigation; Post-discharge transportation assistance; | Inpatient hospital | OUD; AUD; Cocaine UD | No | Methadone; Buprenorphine | Nurse; Social worker/case manager; Multidisciplinary addiction care team; Addiction counselor; Medical doctor | Called upon as part of Addiction consult team | Social worker/case manager | Employed by external organization | SUD treatment providers (unspecified); Other community resources |
| Hu, 2019 [19] | Pre-discharge electronic referral;Post discharge bridge prescription ; Pre-discharge provider list | ED | OUD | No | Buprenorphine | None specified | Work directly in ED/Hospital | Nurse; Medical doctor | Work directly in ED/Hospital | Treatment with MOUD |
| Jennings, 2021 [20] | Pre-discharge schedule appointment | ED | SUD (general); OUD | Yes | Buprenorphine | None specified | Other: | Peer; Medical doctor | Work directly in ED/Hospital | Treatment with MOUD |
| Kaucher, 2020 [21] | General linkage to treatment | ED | OUD | No | Buprenorphine | Nurse practitioner; Physician assistant | Work directly in ED/Hospital | None specified | Work directly in ED/Hospital | Treatment with MOUD |
| Kelly, 2020 [22] | Pre-discharge discuss treatment options; Post discharge bridge prescription; Pre-discharge schedule appointment | ED | OUD | No | Buprenorphine | Nurse; Medical doctor - MD | Work directly in ED/Hospital | Nurse; Social worker/case manager; Medical doctor (MD) | Work directly in ED/Hospital | SUD treatment providers (unspecified); Treatment with MOUD; Primary care providers; Bridge/short term clinic; Other medical and mental health services |
| Kmiec, 2019 [23] | Pre-discharge provider list; Post discharge bridge prescription; Post discharge follow up texts | ED | SUD (general); OUD; AUD; Sedative UD | No | Buprenorphine | Nurse; Medical doctor | Work directly in ED/Hospital | Automated texts + "unspecified" treatment team that replied to unsolicited texts | Offsite | SUD treatment providers (unspecified) |
| Krupski, 2010 [24] | General linkage to treatment | ED | SUD (general); AUD | Yes | None | Addiction counselor, Medical doctor | Work directly in ED/Hospital | Medical doctor | Work directly in ED/Hospital | SUD treatment providers (unspecified) |
| LeSaint, 2020 [25] | Pre-discharge schedule appointment ; Post discharge bridge prescription | ED | OUD | No | Buprenorphine | None specified | Work directly in ED/Hospital | Medical doctor | Work directly in ED/Hospital | Treatment with MOUD |
| McLane, 2020 [26] | Pre-discharge schedule appointment; Post discharge bridge prescription; Pre-discharge electronic referral | ED | OUD | No | Buprenorphine | None specified | Not specified | None specified | Work directly in ED/Hospital | Treatment with MOUD |
| Monico, 2020 [27] | Pre-discharge discuss treatment options; Pre-discharge schedule appointment; Post-discharge follow up calls; Post discharge peer support; Post-discharge care navigation | ED | SUD (general); OUD | Yes | Buprenorphine | Nurse; Peers; “Medical assessment staff" | Work directly in ED/Hospital | Nurse; Peers; Medical doctor | Work directly in ED/Hospital | SUD treatment providers (unspecified); Treatment with MOUD |
| Nordeck, 2018 [28] | General linkage to treatment | Inpatient hospital | OUD; AUD, Cocaine UD | No | Buprenorphine; methadone | Medical doctor | Work directly in ED/Hospital | Medical doctor; Addiction counselor; Social worker/case manager; Nurse | Work directly in ED/Hospital | SUD treatment providers (unspecified), Treatment with MOUD |
| O'Connell, 2020 [29] | Post-discharge transportation assistance; Post discharge peer support | Inpatient hospital | SUD (general) | No | None | None specified | Not specified | Peer (e.g. peer specialist/peer recovery coach) | Work directly in ED/Hospital | SUD treatment providers (unspecified); Support groups |
| Pecoraro, 2012 [30] | Pre-discharge schedule appointment; Post-discharge transportation assistance; Post-discharge follow up calls; | Inpatient hospital | AUD | Yes | None | Nurse; Patient engagement specialists | Called upon as part of Addiction consult team | Patient engagement specialists | Employed by external organization | SUD treatment providers (unspecified); Other community resources |
| Regan, 2021 [31] | Pre-discharge electronic referral ; Post-discharge follow up calls; Pre-discharge schedule appointment; Post discharge bridge prescription | ED | SUD (general); OUD | No | Buprenorphine | None specified | Work directly in ED/Hospital | Medical doctor | Work directly in ED/Hospital | Bridge/short term clinic |
| Rochat, 2004 [32] | Pre-discharge schedule appointment | Inpatient hospital | AUD | No | None | Social worker/case manager; Multidisciplinary addiction care team; Medical doctor | Work directly in ED/Hospital | Social worker/case manager; Multidisciplinary addiction care team; Medical doctor | Work directly in ED/Hospital | SUD treatment providers (unspecified); Residential/inpatient treatment providers |
| Roncero, 2012 [33] | General linkage to treatment | ED | SUD (general) | No | None | Medical doctor | Work directly in ED/Hospital | None specified | Work directly in ED/Hospital | SUD treatment providers (unspecified) |
| Samuels, 2018 [34] | Pre-discharge provider list; Pre-discharge discuss treatment options; Pre-discharge schedule appointment; Post-discharge care navigation; General linkage to treatment | ED | OUD | No | None | None specified | Work directly in ED/Hospital | Peer; Medical doctor | Work directly in ED/Hospital | Treatment with MOUD |
| Schwarz, 2019 [35] | Pre-discharge provide list of programs; Pre-discharge discuss treatment options; General linkage to treatment;Pre-discharge schedule appointment/warm handoff to specific program | ED; Inpatient hospital | AUD | No | None | Addiction counselor | Called upon as part of Addiction consult team | Addiction counselor | Employed by external organization | SUD treatment providers (unspecified) |
| Shanahan, 2010 [36] | Pre-discharge schedule appointment; Pre-discharge discuss treatment options | Inpatient hospital | OUD | No | Methadone | Nurse; Medical doctor | Work directly in ED/Hospital | Nurse | Work directly in ED/Hospital | Treatment with MOUD; Detox program; Residential/inpatient treatment providers; Bridge/short term clinic; Harm reduction services |
| Shaw, 2020 [37] | General linkage to treatment;Pre-discharge provider list Pre-discharge electronic referral; Post-discharge follow up calls | ED | OUD | No | None | None specified | Not specified | Medical doctor; Staff at the treatment clinic | Work directly in ED/Hospital | SUD treatment providers (unspecified) |
| Smith, 2021 [38] | Post discharge follow up calls | Inpatient hospital | OUD; AUD | No | Buprenorphine; Naltrexone | Nurse; Multidisciplinary addiction care team; Medical doctor; Pharmacist | Work directly in ED/Hospital | Pharmacist | Work directly in ED/Hospital | SUD treatment providers (unspecified); Treatment with MOUD; Bridge/short-term clinic |
| Sorensen, 2005 [39] | Post-discharge care navigation | ED; Inpatient hospital | OUD | No | Methadone | Multidisciplinary addiction care team; Research assistant | Work directly in ED/Hospital | Social worker/case manager; Addiction counselor | Work directly in ED/Hospital | Treatment with MOUD; Detox program; Other medical and mental health services; Other community resources |
| Sullivan, 2021 [40] | Pre-discharge discuss treatment options; Pre-discharge schedule appointment | ED | OUD | No | Buprenorphine | None specified | Work directly in ED/Hospital | Peer; Medical doctor | Work directly in ED/Hospital | Bridge/short-term clinic |
| Trowbridge, 2017 [41] | Pre-discharge schedule appointment; Pre-discharge discuss treatment options | Inpatient hospital | OUD | No | Methadone; Buprenorphine; Naltrexone | Multidisciplinary addiction care team; Medical doctor; Nurse practitioner | Work directly in ED/Hospital | Social worker/case manager; Multidisciplinary addiction care team; Medical doctor; Nurse practitioner | Work directly in ED/Hospital | SUD treatment providers (unspecified); Treatment with MOUD; Bridge/short-term clinic |
| Wakeman, 2020a [42] | Pre-discharge discuss treatment options; Pre-discharge schedule appointment; Post discharge bridge prescription | Inpatient hospital | OUD; AUD; Methamphetamine & other stimulant UD; Cocaine UD | No | Methadone; Buprenorphine; Naltrexone | Social worker/case manager; Multidisciplinary addiction care team; Peers; Medical doctor; Nurse practitioner | Work directly in ED/Hospital | Nurse; Social worker/case manager; Peer; Medical doctor; Resource Specialist | Employed by external organization | SUD treatment providers (unspecified); Treatment with MOUD; Bridge/short-term clinic; Residential/inpatient treatment providers; ; Primary care providers |
| Wakeman, 2020b [43] | Pre-discharge schedule appointment; Post-discharge care navigation; Post-discharge follow up calls; | Inpatient hospital | SUD (general) | No | None | Multidisciplinary addiction care team | Not specified | Social worker/case manager; Peer | Work directly in ED/Hospital | SUD treatment providers (unspecified) |
| Watson, 2021 [44] | Post-discharge transportation assistance; Pre-discharge schedule appointment | ED | OUD | No | Yes, unspecified | Multidisciplinary addiction care team | Work directly in ED/Hospital | Social worker/case manager; Multidisciplinary addiction care team; Peer | Called upon to ED/hospital as a consult | SUD treatment providers (unspecified); Treatment with MOUD; Other community resources |
| Whiteside, 2017 [45] | Pre-discharge schedule appointment; Post-discharge follow up calls; Post-discharge care navigation | ED | OUD; Sedative UD | Yes | None | Social worker/case manager; Multidisciplinary addiction care team; Medical doctor; Clinical psychologist | Work directly in ED/Hospital | Multidisciplinary addiction care team; Medical doctor; Research Assistant | Work directly in ED/Hospital | SUD treatment providers (unspecified); Treatment with MOUD; Primary care providers; Other medical and mental health services |

Note: ED=Emergency Department, SUD= Substance Use Disorder, OUD= Opioid Use Disorder, AUD= Alcohol Use Disorder, UD=Use Disorder, MOUD=Medications for Opioid Use Disorder
